# Association Between Preparedness and Response Measures and COVID-19 Incidence and Mortality

**DOI:** 10.1101/2021.02.02.21251013

**Authors:** Christopher T. Lee, Marine Buissonnière, Amanda McClelland, Thomas R. Frieden

## Abstract

The COVID-19 pandemic is the most disruptive global health threat in a century. We analyzed publicly available data on preparedness capacity, COVID-19 incidence and mortality, governance, and testing. Although other analyses have suggested that preparedness assessments do not correlate with effective pandemic response, we found that testing rates correlate with both COVID-19 incidence and mortality and strongly correlated with country preparedness capacity as measured by the Joint External Evaluation (JEE). There is a statistically significant association between preparedness capacities and COVID-19 case incidence and an independent association between governance and COVID-19 case and mortality rates. Legislation, surveillance, and risk communication capacities were associated with lower COVID-19 case incidence and mortality. Preparedness and governance are independently associated with COVID-19 pandemic severity. Preparedness capacities are not sufficient — capacity and governance are both critical to pandemic control. Countries must improve public health systems and implement strong government leadership.

**Article Summary:** Country preparedness capacities and country leadership policy response (governance) are both critically important to control pandemics. Without appropriate policy action, public health preparedness is insufficient for effective pandemic response.

## Background and Introduction

The COVID-19 pandemic has been the most disruptive global health threat in a century and, as of this writing, continues to accelerate, with more than 80 million diagnosed cases and more than 1.8 million recorded deaths worldwide by the end of 2020. The United States reported more than 20 million cases (with at least 3 times as many estimated to have been infected), with nearly 350,000 deaths – a rate of one death every 30 seconds as of mid-December 2020. The introduction of highly effective vaccines will not markedly change the trajectory of the pandemic for several more months, until large portions of the population can receive vaccination.

The WHO International Health Regulations (2005) were revised after the 2003 SARS outbreak with the goal of making the world better prepared to address infectious disease threats. The Joint External Evaluation (JEE) process, a voluntary, transparent, and objective assessment of each country’s level of preparedness, measures capacity in 19 key areas and identifies specific gaps. The JEE is an external assessment of progress meeting IHR commitments and creating National Action Plans for Health Security (NAPHS).^1,2^ Since 2017, JEEs have been conducted in 127 of 194 WHO Member States.

In addition to periodic JEE assessments, countries report progress on meeting IHR (2005) requirements using the State Party Annual Reporting (SPAR) tool. Early in the pandemic, WHO analyzed SPAR data to anticipate country capacities for managing COVID-19 and determined that half of all countries reviewed had strong operational readiness capacities in place, suggesting that that an effective response to potential health emergencies, including to the COVID-19 pandemic, could be implemented.^3^ Thus, it would seem that results of JEE assessments should be related to the level of success of COVID-19 response. However, several other analyses did not find any correlation between JEE scores and COVID-19 case and death rates, with these analyses including both the JEE and the Global Health Security Index.^4,5,6^ There is a perception that the JEE has not correlated with the effectiveness of country response to the pandemic.^7^

In addition to country preparedness capacities, the COVID-19 pandemic has shown that a country’s policy response – driven by the highest levels of leadership — is also critically important to manage the pandemic.^8,9^ That is, country preparedness capacities as measured by JEE assessments, as well as the actions that governments take — broadly described as governance — both may be critical to reduce COVID-19 case incidence and mortality. We hypothesized that higher levels of country preparedness capacities as measured by the JEE were independently associated with COVID-19 case incidence and mortality, and that a separate measure of country response — governance — was also associated with COVID-19 case incidence and mortality.

## Methods

### Data sources

We analyzed publicly available data from WHO Joint External Evaluation mission reports; data were available for 94 countries.^10^ We used an aggregate measure of JEE scores using a methodology applied elsewhere that averages the 19 technical area scores from the JEE.^2^ We compared JEE score data to other capacity indices, including the Global Health Security Index (GHSI)^11^ and Universal Health Coverage (UHC) effective coverage ratings.^12^ We found substantial covariance among these three measures, and focused on JEE scores for capacity measurement; the correlations among these measures have been published elsewhere.^4,5,6^

For COVID-19 case incidence and mortality rate (per million), we used data from the European Centre for Disease Prevention and Control, as of October 27, 2020.^13^ Both case and death data were right-tailed, so we applied a logarithmic transformation to normalize the data to make suitable for use as an outcome measure. We used data from Our World in Data for testing data,^14^ available daily for 49 countries that had also conducted a JEE. To develop a static measure of country testing response rates, we used the cumulative testing rate (tests per thousand) for which data were available. These data were also right-tailed, so we applied a logarithmic transformation to normalize the data.

We used the COVID-19 Global Response Index, developed by *Foreign Policy*,^8^ to assess effective national policy responses in 36 countries. There were 16 countries with both JEE scores and testing data available. The Global Response Index uses publicly available data to track public health directives, financial responses, and fact-based public communications. The Index was originally published on August 1, 2020, and scores were updated on October 1; we use the latter data for this analysis.

### Data analysis

We conducted exploratory data analysis for capacity data (JEE score, GHSI, UHC effective coverage), capability data (Global Response Index, confidence in government), and outcomes data (COVID-19 log-transformed incidence and mortality). We examined the covariance structure for all variables in order to inform model selection and reduce statistical confounding. We conducted stratified analyses to further explore the association between JEE score and COVID-19 case incidence, stratifying by quartile of COVID-19 testing rate, to determine whether the direction of association changed after stratifying by testing rate, as JEE scores were highly correlated with testing rates.

After we observed a linear association between covariates of interest and outcomes, we performed multiple linear regression analysis using the log-transformed outcome variables. We repeated these analyses using scores for six JEE technical areas of particular relevance to the COVID-19 pandemic: legislation, policy, and finance (P1); laboratory systems (D1); surveillance (D2); emergency operations (R2); risk communication (R5); and points of entry (POE). We used Akaike’s Information Criterion (AIC) to measure model fit and parsimony to inform the final model selection.

## Results

### Association between country capacity scores, testing rates, and COVID-19 incidence and mortality rates

Measures of COVID-19 incidence and mortality (per million) were strongly correlated (Table 1). Among 49 countries with available testing data, testing rate (logarithm of total tests conducted per thousand population) was moderately correlated with both COVID-19 incidence and mortality (Pearson’s *r* = 0.58 and 0.62, respectively) and more strongly correlated with country capacity scores from the JEE *(r =* 0.76). Measures of health systems capacity by the GHSI and UHC effective coverage were strongly correlated with JEE scores and did not meaningfully explain additional variance in measures of COVID-19 incidence and mortality.

**Table 1:** Correlation Matrix (Pearson’s *r*) of Capacity, Capability, and Outcome Variables — Global as of October 27, 2020

|  | Log Cases per Million* | Log Deaths per Million* | Log Test Rate per Thousand* | Joint External Evaluation Ready Score | Global Health Security Index | Universal Health Coverage (Effective Coverage) | Foreign Policy Response Index |
| --- | --- | --- | --- | --- | --- | --- | --- |
| Log Cases per Million* | 1.0 |  |  |  |  |  |  |
| Log Deaths per Million* | 0.97 | 1.0 |  |  |  |  |  |
| Log Test Rate per Thousand* | 0.58 | 0.62 | 1.0 |  |  |  |  |
| Joint External Evaluation Ready Score | 0.21 | 0.30 | 0.76 | 1.0 |  |  |  |
| Global Health Security Index | 0.37 | 0.49 | 0.71 | 0.85 | 1.0 |  |  |
| Universal Health Coverage (Effective Coverage) | 0.20 | 0.26 | 0.72 | 0.95 | 0.80 | 1.0 |  |
| Foreign Policy Response Index* | -0.70 | -0.70 | -0.46 | -0.36 | -0.56 | -0.35 | 1.0 |

\*As of October 27, 2020

Among 16 countries with both Global Response Index and JEE data available, there was a negative and moderately strong correlation between the Global Response Index and COVID-19 measures (*r* = –0.70 for both incidence and mortality) and a positive and weak correlation between the JEE score and COVID-19 incidence (*r* = 0.21) and mortality (*r* = 0.28). For these 16 countries, preparedness as measured by JEE and governance as measured by the Global

Response Index were independent (not correlated) (*r* = –0.36). Some countries with better preparedness as measured by the JEE, such as the United States, with a JEE score 87 out of 100, had low Foreign Policy response indices (19 in August 2020 and 39 in October 2020), whereas Senegal had low preparedness (JEE score 45) but among the highest Foreign Policy response indices (89 in August 2020 and 83 in October 2020) (Table 2).

**Table 2:** Joint External Evaluation Scores (Preparedness) and Foreign Policy Response Indices — 16 Countries

| **Country** | **Joint External**<br>**Evaluation Score** | **Foreign Policy**<br>**Response Index**<br>**(August 2020)** | **Foreign Policy**<br>**Response Index**<br>**(October 2020)** |
| --- | --- | --- | --- |
| **Australia** | 92 | 62 | 93 |
| **Belgium** | 85 | 67 | 47 |
| **Canada** | 93 | 47 | 76 |
| **Switzerland** | 89 | 41 | 45 |
| **Ethiopia** | 52 | 40 | 88 |
| **Finland** | 86 | 64 | 57 |
| **Ghana** | 45 | 61 | 92 |
| **Indonesia** | 64 | 21 | 69 |
| **Japan** | 92 | 47 | 97 |
| **Kenya** | 50 | 62 | 94 |
| **Republic of Korea** | 92 | 48 | 51 |
| **New Zealand** | 89 | 100 | 100 |
| **Saudi Arabia** | 76 | 80 | 76 |
| **Senegal** | 45 | 89 | 83 |
| **United States of America** | 87 | 19 | 39 |
| **South Africa** | 62 | 65 | 69 |

Because testing rates were correlated with JEE scores as well as COVID-19 incidence and mortality measures, we stratified the association between JEE scores and COVID-19 incidence (Figure 1) based on testing quartile. Post-stratification, the direction of the association between JEE scores and COVID-19 incidence reversed for 3 of 4 testing quartiles, indicating confounding of the association between JEE scores and COVID-19 incidence by testing rates.

**Figure 1:**
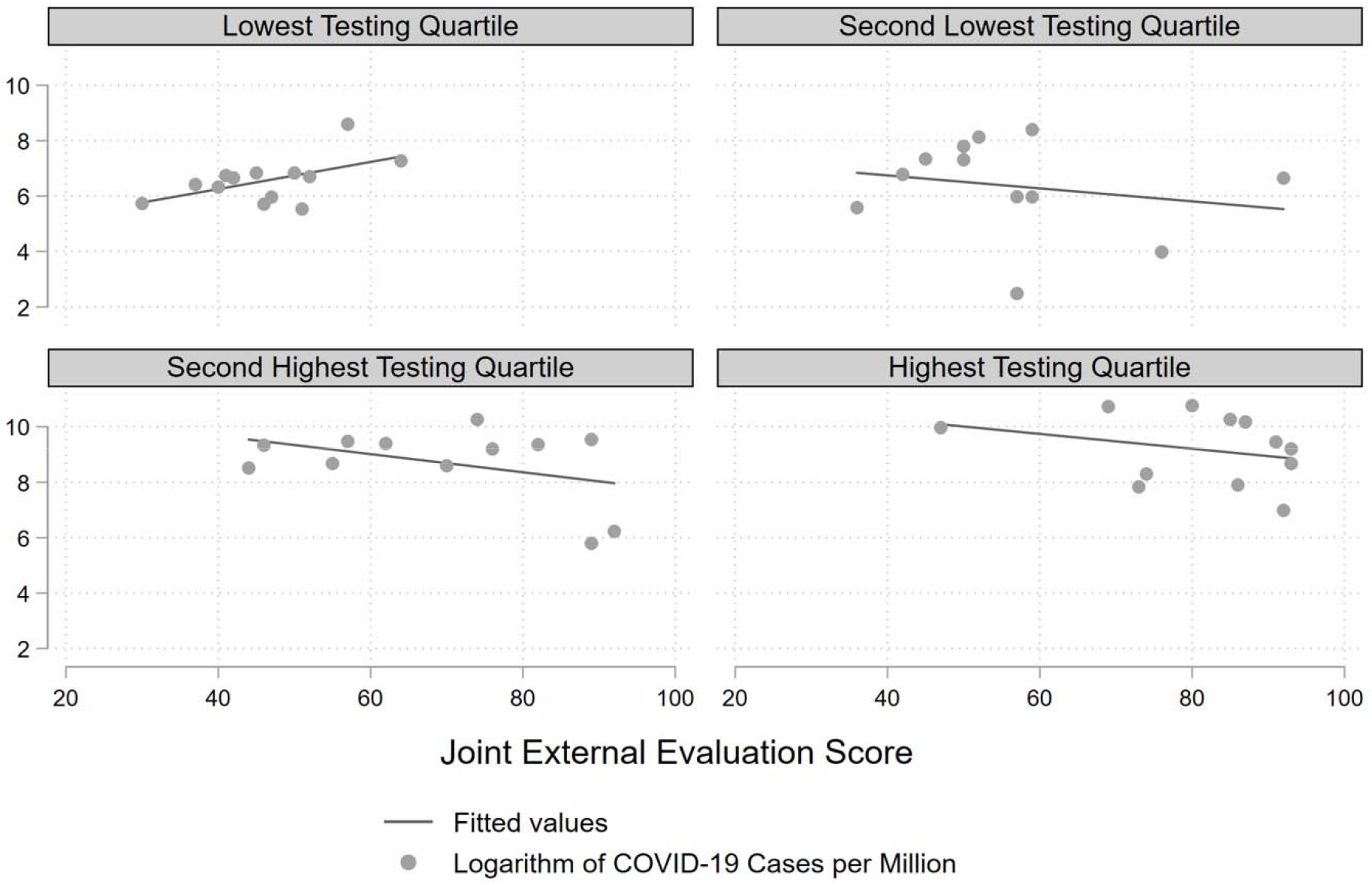
Logarithm of Country COVID-19 Cases per Million and Joint External Evaluation Scores, Stratified by Testing Quartile — 49 Countries as of October 27, 2020

In a multiple linear regression model, adjusting for testing rates, JEE score and Foreign Policy response index were independently associated with lower COVID-19 case incidence (Table 3). For COVID-19 incidence, the variance explained by JEE score s (*P* = 0.037) was independent of that of the Foreign Policy response index (*P* = 0.009), and the model containing both variables had the best fit as measured by AIC. A similar relationship was observed for COVID-19 mortality (Table 4), in which both JEE scores and Foreign Policy response index (*P* = 0.015) were independently negatively associated with COVID-19 log mortality, although the JEE score association with COVID-19 mortality did not reach statistical significance (*P* = 0.129).

**Table 3:**
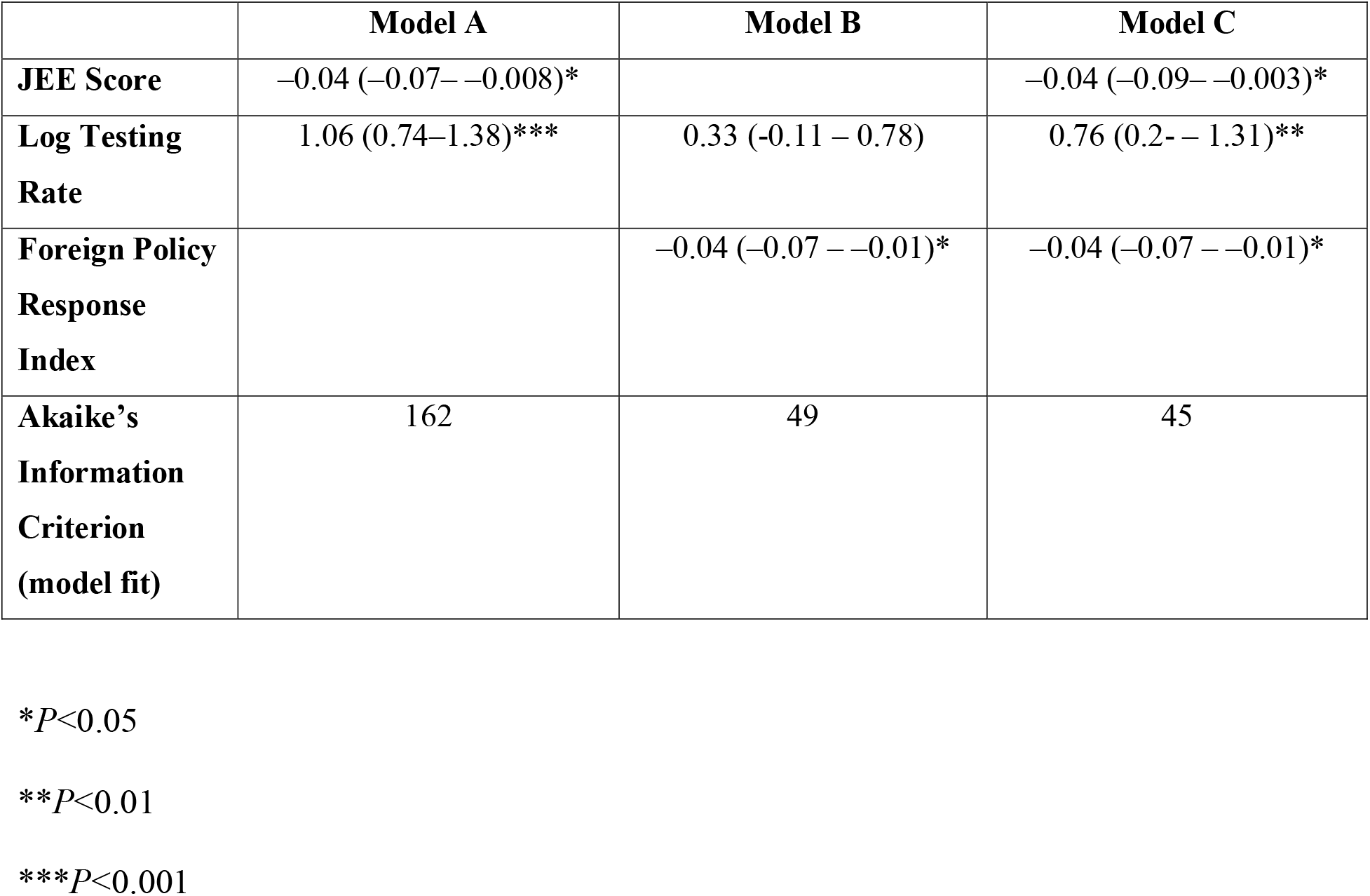
Multiple Linear Regression of COVID-19 Cases per Million (logarithm) and Explanatory Variables, Adjusted by Testing Rate (logarithm)

**Table 4:**
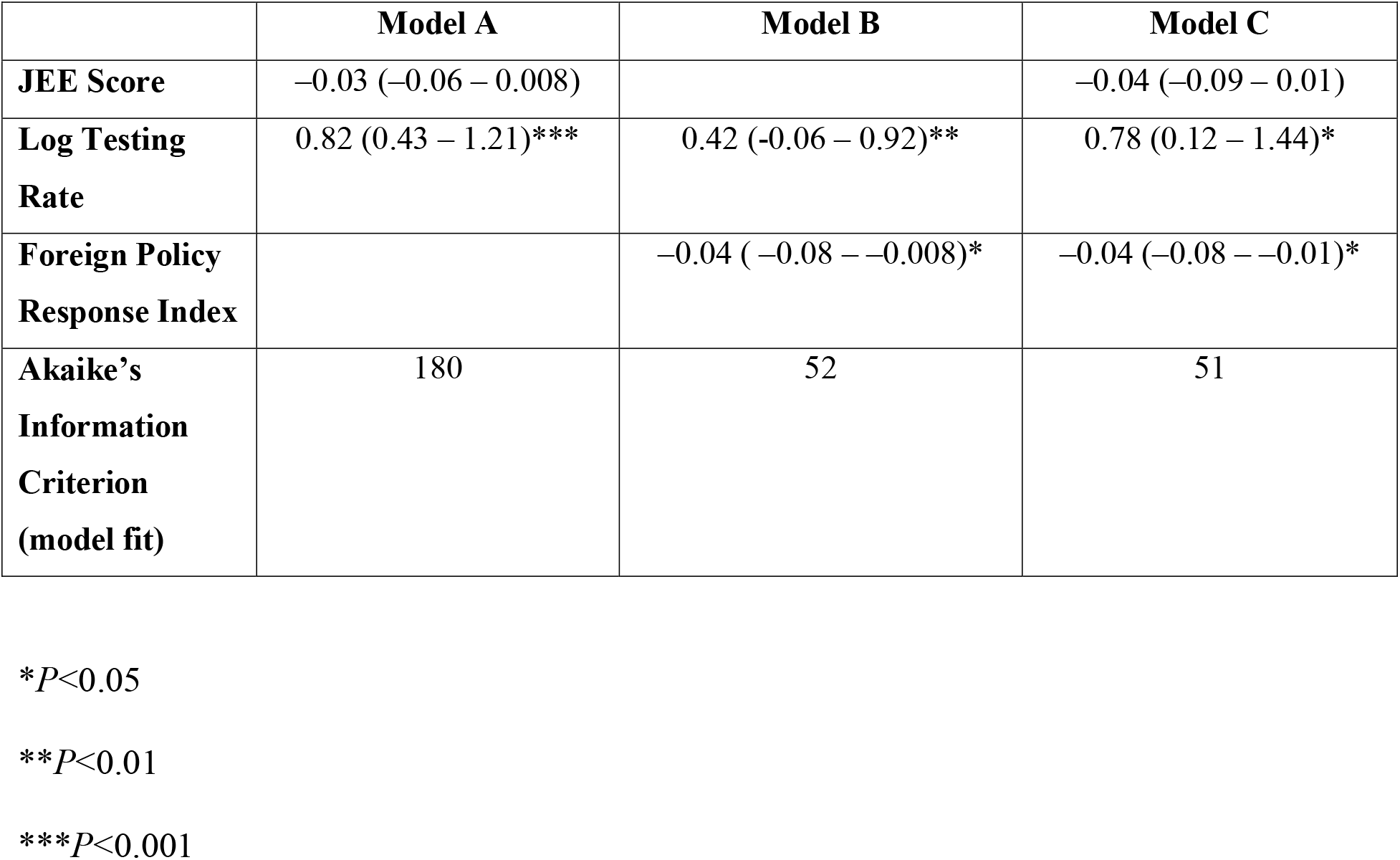
Multiple Linear Regression of COVID-19 Deaths per Million (logarithm) and Explanatory Variables, Adjusted by Testing Rate (logarithm)

When the linear regression model was replicated for specific JEE technical areas, we found that legislation (*P* = 0.001), surveillance (*P* = 0.005), and risk communication (*P* = 0.01) were significantly negatively associated with COVID-19 incidence (Table 5). Legislation (*P* = 0.006), surveillance (*P* = 0.01), and risk communication (*P* = 0.008) were also significantly negatively associated with COVID-19 mortality. The other three technical areas we analyzed (laboratory, emergency operations, and points of entry) were not significantly associated with either COVID-19 incidence or mortality.

**Table 5:**
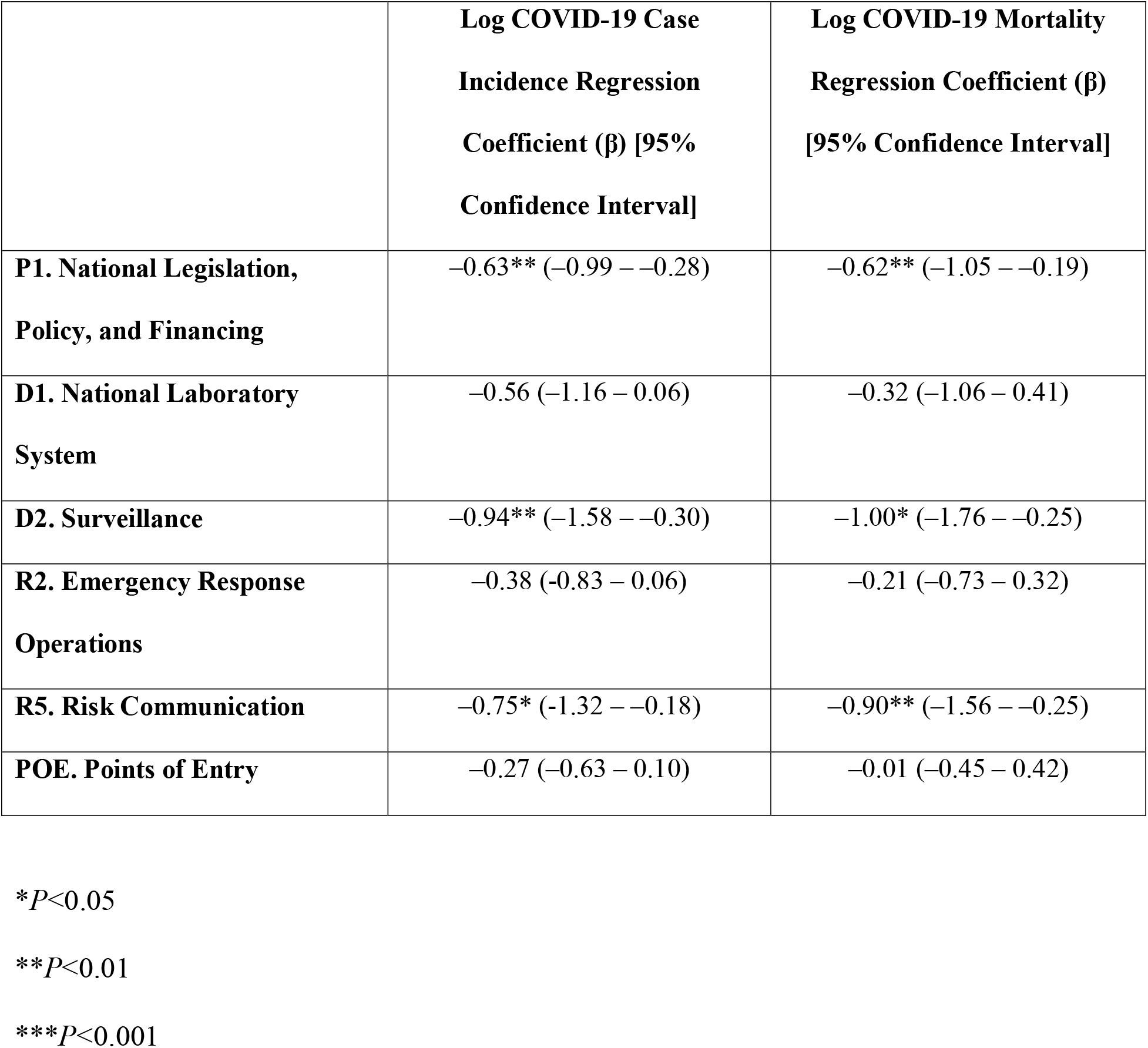
Multiple Linear Regression of COVID-19 Incidence and Deaths per Million (logarithm) and Selected Joint External Evaluation Technical Areas, Adjusted by Testing Rate (logarithm) (n = 49 countries)

## Discussion

In this analysis, we found that, after adjusting for testing rate, there is a statistically significant association between preparedness capacities as measured by the JEE and COVID-19 case incidence. We also found an independent association between the strength of the government policy response and COVID-19 case incidence and mortality rates. Taken together, we find that preparedness capacities and governance are independently associated with the severity of the COVID-19 pandemic at the country level. Countries must invest in preparedness capacities as well as implement strong government leadership and utilize evidence-based policies during crises.

The association between preparedness scores and COVID-19 case incidence was confounded by testing rates. Higher testing rates were observed in countries with higher JEE scores, and were also associated with higher numbers of reported cases. When we stratified the data by testing rate, we found the direction of association between preparedness scores and COVID-19 case incidence to reverse, which confirmed the presence of confounding: countries with stronger public health systems had higher JEE scores, more tests per capita, and a higher reported incidence rate, but were likely to have had a less severe pandemic. Countries with higher testing capacities are likely to identify more cases, either because of higher disease prevalence or a better rate of disease detection,^15^ which is critical for control of COVID-19. Conversely, countries with lower testing rates are likely to underestimate the burden of disease.

### Building Preparedness Capacities

Previous analyses have not found an association between preparedness measures and COVID-19 case counts or incidence, and have questioned the value of the JEE process.^4,5,6^ We find that preparedness scores do in fact correlate with pandemic severity, but that this association is only apparent after adjusting for testing rates, which we identified as a confounding variable. The JEE is a useful tool to measure country progress on achieving the IHR (2005) commitments and provide the necessary planning framework that can be costed and implemented into a fully functional action plan.^1^ Critical preparedness capacities include implementation of enabling legislation, sufficient sustainable financing, coordination among sectors, frameworks for effective infection prevention and control, and core capacities of surveillance, laboratory, workforce, risk communication, and emergency response operations.

We found that legislation, surveillance, and risk communication were significantly associated with lower COVID-19 incidence and mortality. These preparedness capacities represent important policies and financing for implementation of the IHR (2005) as well as the capacity to detect events, report them, and communicate them to the public. Laboratory systems were not significantly associated with COVID-19 incidence or mortality, which is likely an artifact of confounding as analyses were adjusted by testing rates, which were strongly correlated with laboratory capacity scores.

The JEE tool may benefit from revisions based on lessons learned from the COVID-19 pandemic, particularly in developing a new technical area for safer health facilities, as we have proposed elsewhere.^16^ Similarly, the JEE does not rigorously measure country health systems’ ability to provide essential services for both primary and advanced respiratory care. However, the JEE process and subsequent planning and implementation processes remain a critical component to objectively measure and improve country preparedness capacities and should be refined rather than rejected.

Effective planning requires program management and financing expertise, a strong domestic commitment to financing and implementation, and advocacy for increased long-term domestic financing, as well as accountability for transparently making progress on preventing epidemics to make populations safer and meet global commitments.^2^ Substantial investments are required globally to increase preparedness, ranging from an up-front two-year investment of $20 billion to $30 billion and ensuing annual investments of $5 billion to $10 billion (for a ten-year total of $60 billion to $110 billion),^17^ to as much as $35-40 billion ($5 per person worldwide) per year for the next decade.^18^

The 2014-2016 West Africa Ebola epidemic demonstrated the ongoing cycle of panic and neglect that surrounds public health emergencies.^19^ As the memories of one outbreak or epidemic fade, complacency sets in and commitments to invest in preparedness are set aside, resulting in continued vulnerabilities and eventually crisis when the next outbreak inevitably occurs. In the past two decades, there have been six outbreaks that reached pandemic thresholds, and many others achieved epidemic status but were contained before there was uncontrolled global spread.^20^

The cost of not being prepared is enormous. SARS, H1N1 influenza, and Ebola each cost the global economy between $40 billion and $55 billion in response costs and indirect costs due to lost livelihoods and lives as a result of essential service disruptions. COVID-19 has already cost the world an estimated $20 trillion in direct spending and reduced productivity^21^ — a quarter of one year’s total global GDP — and the pandemic is far from over.

### Governance

The JEE measures country preparedness capacities, but it does not assess government leadership in a crisis. To assess this, we used the Foreign Policy COVID-19 Global Response Index, which tracks country leadership response in critical policy areas, including public health directives, financial responses, and fact-based public communications.^8^ Capacity and governance are complementary, and both are important to a country’s preparedness to effectively address health threats. We found that preparedness capacities as measured by the JEE and governance as measured by the COVID-19 Global Response Index were not correlated — countries with high preparedness levels could have poor responses and vice versa — and preparedness and response independently accounted for variance in COVID-19 case incidence. Governments with strong preparedness capacities also need effective government leadership during public health emergencies, and conversely, strong government responses might compensate in part when preparedness capacities are sub-optimal.

The IHR Monitoring Evaluation Framework has two components that help with measurement of governance – simulation exercises and after-action reviews — both of which are functional assessments of how systems would be expected to or actually did perform.^22^ Simulation exercises attempt to assess capacity to respond to future incidents and help countries plan and allocate resources, and can be performed periodically — particularly when there is a change in government leadership — to pressure-test the coordination mechanisms and prepare leaders. After-action reviews are critical measures of governance and provide an accurate analysis of ongoing or past events to help countries learn from experience. Countries should conduct routine assessments of public health responses to prioritize areas for capacity development using the after-action process and periodically conduct simulation exercises for high-risk events to prepare for and test government coordination mechanisms.

The WHO Thirteenth General Programme of Work (GPW 13) “triple billion” metrics — three targets to promote health, keep the world safe, and serve the vulnerable — aim by 2023 to have one billion more people benefit from universal health coverage, be better protected from health emergencies, and enjoy better health and well-being.^23^ The health emergencies billion component measures both dimensions: capacities as measured by SPAR, and governance capabilities as measured by timeliness to detect, notify, and respond.^24^ Countries should use routine assessment tools including the SPAR and JEE to identify gaps for capacity development and implement data systems, including event management systems, to routinely measure the timeliness of detection, notification, and response to serious public health events. Timeliness of detection, notification, and response can be useful to identify response bottlenecks for performance management, maintain mutual accountability between Member States, and longitudinally monitor country detection and response capacities as well as their commitment to the IHR (2005).

### Limitations

This analysis is subject to several limitations. The cross-country analyses performed might not capture the many complex dynamics of a specific disease, but the measures being tested are generally available only at the national level. There were limited data available on testing (only 49 countries analyzed had completed a JEE) and on policy response (index data were available for only 16 countries with a completed JEE). Other sophisticated modeling efforts reviewed a number of factors to identify predictors of COVID-19; our hypothesis was to test the role of capacities and governance rather than the etiological dynamics of COVID-19 or other infectious diseases, so we do not provide analysis of the predictive effect capacity or governance as defined in this paper. Last, JEEs are only conducted every 4–5 years and the process began in 2017; it is possible that country capacities have changed since the time the JEE was conducted.

## Conclusion

The COVID-19 pandemic has shown that both country preparedness capacities, as primarily measured by WHO Joint External Evaluation assessments, and country leadership policy response (governance) are critically important for pandemic control to reduce case incidence and mortality. Countries and global financial support initiatives need to sustainably invest in preparedness to avoid yet another cycle of panic and neglect. During emergencies, governments need to use science and evidence to inform policy and need to communicate effectively with their populations and issue clear and consistent guidance.

## Data Availability

All data referenced in this article are publicly available as noted in the text.

## Acknowledgments

Resolve to Save Lives, an Initiative of Vital Strategies, is funded by Bloomberg Philanthropies, the Bill & Melinda Gates Foundation, and Gates Philanthropy Partners, which is funded with support from the Chan Zuckerberg Foundation. The authors wish to thank Drew Blakeman for assistance with manuscript preparation.

## Author Bio

Dr. Christopher T. Lee is a medical epidemiologist and is the Director of Global Epidemic Preparedness and Response at Resolve to Save Lives, an Initiative of Vital Strategies. Dr. Lee’s team provides direct technical assistance to countries on strengthening preparedness capacity development, strengthening program management for the accountable and effective use of health security funds, strengthening national and global protection of health care workers, and implementing measures to improve COVID-19 surveillance, case investigation, and contact tracing. His research work interests include infectious disease epidemiology, causes and consequences of forced migration and homelessness, and policy impact assessment affecting persons experiencing homelessness or displacement.

